# COVID-19 Incidence and Hospitalization During the Delta Surge Were Inversely Related to Vaccination Coverage Among the Most Populous U.S. Counties

**DOI:** 10.1101/2021.08.17.21262195

**Authors:** Jeffrey E. Harris

## Abstract

We tested whether COVID-19 incidence and hospitalization rates during the Delta variant-related surge were inversely related to vaccination coverage among the 112 most populous counties in the United States, together comprising 44 percent of the country’s total population. We measured vaccination coverage as the percent of the county population fully vaccinated as of July 15, 2021. We measured COVID-19 incidence as the number of confirmed cases per 100,000 population during the 14-day period ending August 12, 2021 and hospitalization rates as the number of confirmed COVID-19 admissions per 100,000 population during the same 14-day period. In log-linear regression models, a 10-percentage-point increase in vaccination coverage was associated with a 28.3% decrease in COVID-19 incidence (95% confidence interval, 16.8 - 39.7%), a 44.9 percent decrease in the rate of COVID-19 hospitalization (95% CI, 28.8 - 61.0%), and a 16.6% decrease in COVID-19 hospitalizations per 100 cases (95% CI, 8.4 - 24.8%). Inclusion of demographic covariables, as well as county-specific diabetes prevalence, did not weaken the observed inverse relationship with vaccination coverage. A significant inverse relationship between vaccination coverage and COVID-19 deaths per 100,000 during August 20 – September 16 was also observed. The cumulative incidence of COVID-19 through June 30, 2021, a potential indicator of acquired immunity due to past infection, had no significant relation to subsequent case incidence or hospitalization rates in August. Higher vaccination coverage was associated not only with significantly lower COVID-19 incidence during the Delta surge, but also significantly less severe cases of the disease.

## Introduction

By the second week of July 2021, the fast-spreading Delta variant had been detected in more than 99 percent of all SARS-CoV-2 viral isolates reported in the United States.^1^ While the Delta-driven surge in COVID-19 cases in the U.S. initially appeared to have been concentrated in places with relatively low vaccination rates,^2-4^ by early August there were reports of emerging hot spots in highly vaccinated parts of the country.^5^ By mid-August, breakthrough infections in fully vaccinated individuals,^6^ in part the result of a diminution over time in vaccine effectiveness,^7 8^ had risen in some places to as high as 30 percent of all reported cases.^9^ Fully vaccinated individuals, once infected with the Delta variant, were found to be capable of transmitting their infections to others,^10^ though their viral load and duration of infectivity were found to be lower than that of unvaccinated infected individuals.^11^

In view of these developments, we conducted an observational, cross-sectional analysis of the relation between vaccination coverage and COVID-19 disease rates among counties in the United States during the Delta-driven surge. Specifically, we tested whether COVID-19 incidence and hospitalization rates during the two weeks ending August 12 were inversely related to the percentage of the population fully vaccinated by mid-July 2021. To avoid comparing small rural counties with large urban centers, we concentrated on the 112 largest counties, each with a population over 600,000, and together with a combined total population of 147 million persons, or about 44 percent of the entire U.S. population.

## Data and Methods

### Data

#### Principal Analyses

Our data derive principally from the *COVID-19 Community Profile Report* maintained at *healthdata*.*gov*.^12^ The *Counties* tab in the spreadsheet for 8/12/2021 gave the incidence of COVID-19 cases per 100,000 during the most recent and the previous 7-day periods, from which we calculated the *14-day cumulative incidence*. The spreadsheets for 8/5/2021 and 8/12/2021 gave the numbers of confirmed COVID-19 hospitalizations for the two previous 7-day periods, from which we computed county-specific *14-day hospital admission rates* per 100,000. We also computed the number of *COVID-19 hospital admissions per 100 cases*, which we defined as 100 times the hospital admission rate divided by the incidence rate.

We similarly relied on the *Counties* tab in the spreadsheet for 7/15/2021 to extract the county-specific percentage of the population fully vaccinated as of that date. Since vaccination coverage for Texas was omitted from the *Community Profile Report*, we supplemented our database with state-specific data compiled by the *Democrat and Chronicle* as of 7/14/21.^13^

These sources, taken together, provided us with one independent variable – the vaccination coverage in each county as of mid-July – and three dependent variables – 14-day COVID-19 incidence, 14-day COVID-19 hospital admission rates, and COVID-19 hospital admissions per 100 cases – in each county for the period ending August 12. These variables together served as the basis of our principal regression analyses, described below.

#### Ancillary Analyses

In a series of ancillary analyses, we considered two additional dependent variables: (1) the *test positivity rate*, defined as the 14-day incidence of COVID-19 divided by the total number of polymerase chain reaction (PCR) diagnostic tests for COVID-19 during the same 14-day period ending 8/12/21; and (2) the COVID-19 *death rate*, defined as the cumulative number of deaths from COVID-19 per 100,000 population recorded during the 4-week interval from 8/20 through 9/14/21. The data underlying the test positivity rate were derived from the 8/12/21 spreadsheet, while the data underlying the death rate were derived from the 8/19/21 and 9/14/21 spreadsheets of the *COVID-19 Community Profile Report*.

In our ancillary analyses, we also considered the following additional independent variables: (1) the cumulative number of confirmed COVID-19 cases per 100 population in each county as of 6/20/2021; (2) the fraction of the county population aged 65 years or more; (3) the fraction of the county population reported as non-Hispanic black; (4) the fraction of the county population reported as Hispanic; (5) the Center for Disease Control’s (CDC’s) Social Vulnerability Index (SVI) for each county;^14^ and (6) the prevalence of diabetes among persons aged 20 year or more in each county in 2018. Cumulative confirmed cases through 6/20/21 were computed from the New York Times and New York City Department of Public Health databases.^15 16^ Diabetes prevalence was derived from the CDC’s *Diabetes Atlas*.^17^ The remaining independent variables were derived from the 8/12/21 spreadsheet of the *COVID-19 Community Profile Report*.

### Statistical Methods

#### Principal Analyses

In our principal analyses, we identified 112 counties with population at last 600,000. These counties are mapped in Fig. A1 in Appendix A and enumerated in the accompanying legend. While these 112 counties represented only 3.4% of the total of 3,272 counties enumerated in the *COVID-19 Community Profile Report*, their combined population of 147 million represented 44.4% of the total U.S. population of 331 million.

We first conducted a descriptive analysis of the data. To that end, we divided our study sample of 112 counties into 56 counties in the lower half and 56 counts in the upper half of the distribution of vaccination coverage. We computed the means for each of the three dependent variables in both the lower and upper halves and then relied on the t-test based upon unequal variances to assess differences in group means.

We then conducted a cross-sectional regression analysis of the sample of 112 counties, where each county constituted a distinct observation. We employed ordinary least squares (OLS) to estimate the parameters (*α, β*) of the log-linear model log *Y* = *α* + *β X*, where *Y* is the dependent variable of interest in each county (that is, COVID-19 incidence, COVID-19 hospitalization rate, or the hospitalization-case ratio) and *X* represents the corresponding vaccination coverage in that county. In our results below, we report these estimates as Model 1. We also estimated the same log-linear model by population-weighted least squares (reported as Model 2). We further estimated the model log *Y* = *α* + *β X* + *μ*_*FL*_ + *μ*_*TX*_, where *μ*_*FL*_ and *μ*_*TX*_, respectively, are binary parameters indicating whether the county was one of the 10 located in Florida or one of the 11 located in Texas (Model 3). We specifically focused counties in these two large, populous states as they were reported to have especially high rates of infection and hospitalization during the Delta variant-driven surge.^18-21^

#### Ancillary Analyses

We conducted several ancillary analyses to test the robustness of our principal findings. First, we re-estimated the regressions in Models 1 through 3 with a larger, alternative database of 138 counties with population 500,000 or more. Second, utilizing the original database of 112 counties, we re-estimated the regressions in Models 1–3 on two alternative dependent variables: the test positivity rate; and the death rate. Third, utilizing the original database of 112 counties, we estimated an expanded model (Model 4) with the specification log *Y* = *α* + *β X* + *γZ*, where the covariate *Z* represented any one of the six additional independent variables enumerated above.

Table A1 in the Appendix enumerates the 26 additional counties included in our alternative database of 138 counties. Table A2 in the Appendix displays the summary statistics for all variables utilized in our regression models on the main 112-county database.

## Results

### Principal Analyses

The median vaccination coverage across all 112 counties was 49.95 percent. Thus, the lower half of the distribution consisted of 56 counties with a vaccination coverage below 49.95 percent, while the upper half consisted of 56 counties with a vaccination coverage equal to at least 49.95 percent. Table 1 gives the mean values of the independent variable and the three dependent variables for the lower and upper halves of the sample. The mean coverage of the lower half of the distribution was 42.61 percent, while the mean coverage of the upper half was 57.37 percent. The mean COVID-19 incidence per 100,000 was 543.8 per 100,000 in the lower half and 280.6 per 100,000 in the upper half (p < 0.0001 in a t-test of group means with unequal variances). The mean COVID-19 hospital admission rate per 100,000 was 55.37 in lower half and 20.48 in the upper half (p < 0.0001). The mean number of COVID-19 hospital admissions per 100 cases was 8.96 in the lower half and 7.06 in the upper half (p = 0.0037).

**Table 1.**
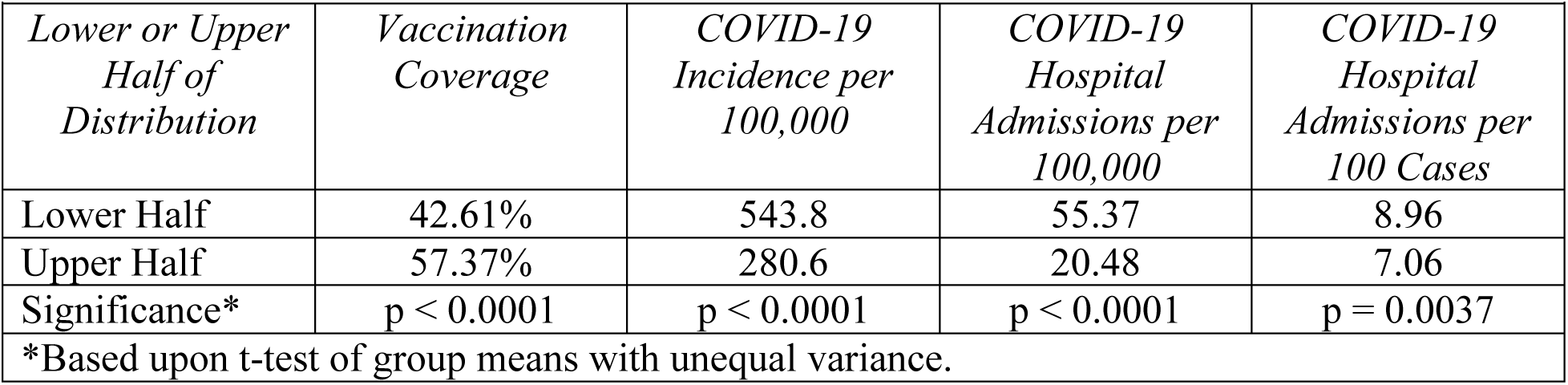
Mean Values for the Lower and Upper Halves of the Vaccination Coverage Distribution.

Table 1 below demonstrates significant absolute differences between the lower and upper halves in COVID-19 incidence, COVID-19 hospital admission rates, and the number of COVID-19 hospital admissions per 100 cases. What’s more, the relative difference in hospital admissions (55.37 / 20.48 = 2.70) is considerably greater than the relative difference in case incidence (543.8 / 280.6 = 1.94), a finding that points to a marked increase in case severity among low-coverage counties. This conclusion is supported by the significant difference between the two halves in the admission-case ratio.

Fig. 1 below displays a two-way scatterplot of COVID-19 incidence versus vaccination coverage in each of the 112 counties. While there is substantial scatter, an inverse relationship is nonetheless evident. The most populous counties in Florida – including Miami-Dade, Palm Beach, Hillsborough (including the city of Tampa), Broward (including Fort Lauderdale), Orange (including Orlando), Duval (including Jacksonville), and others – display notable clustering that suggests a shared determinant. This clustering is not as evident for Texas. While the data points for the interior counties of Bexar County (home to San Antonio) and Harris County (home to Houston) appear relatively close to each other, the cyan data point for the border county of El Paso TX, is situated at the bottom of the plot of Fig. 1.

**Fig. 1.**
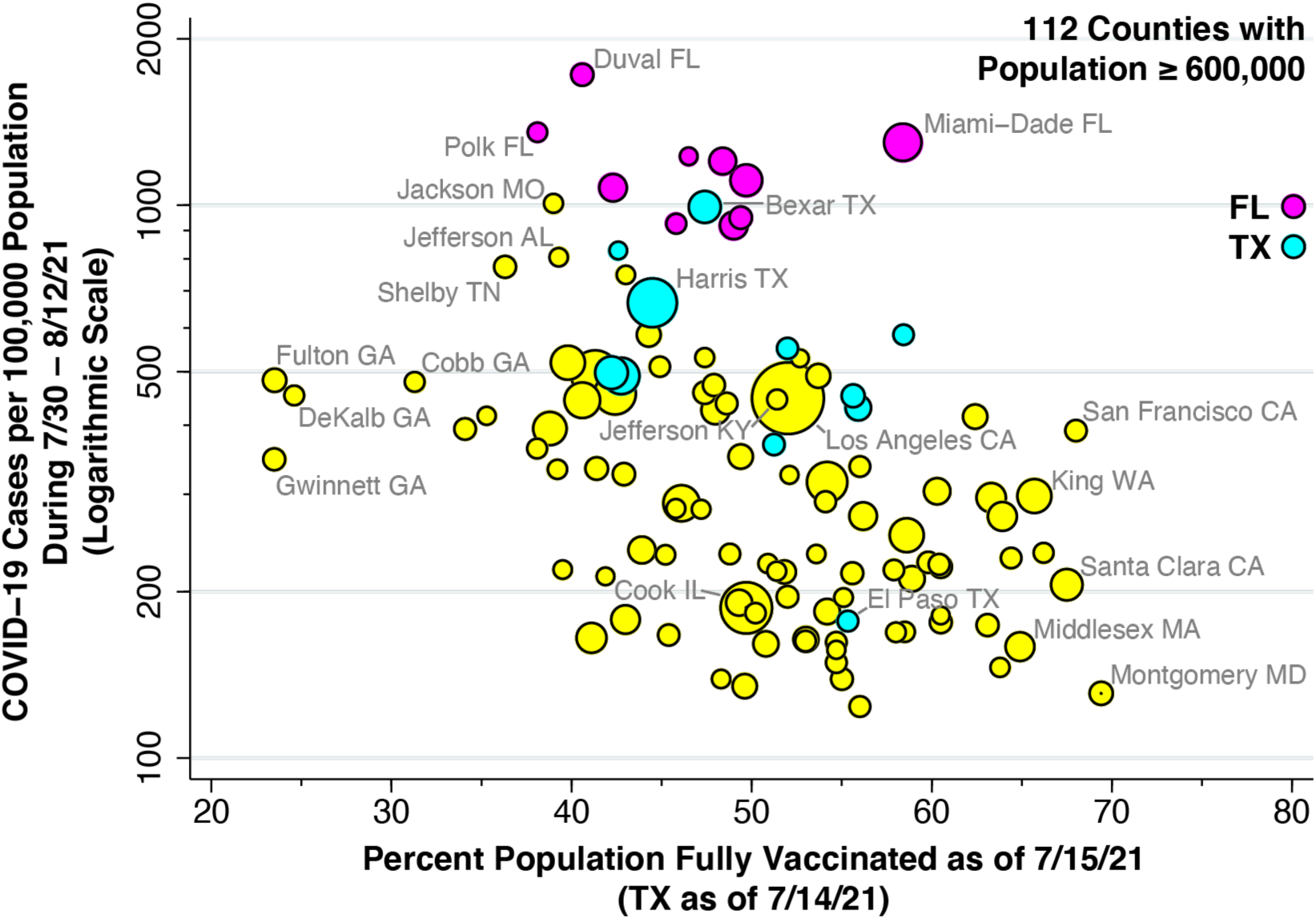
COVID-19 Incidence During 7/30 – 8/12/2021 Versus Vaccination Coverage as of 7/15/2021 in 112 U.S. Counties with Population ≥ 600,000. COVID-19 incidence is measured on a logarithmic scale as confirmed cases per 100,000 population. Vaccination coverage is measured as percent of population fully vaccinated. Vaccination coverage data for 11 Texas counties as of 7/14/2021. Florida counties highlighted in magenta. Texas counties highlighted in cyan. Size of data point proportional to county population.

Fig. 2 below plots hospital admission rates versus vaccination coverage. Again, an inverse relationship is evident. At one extreme, we observe low-vaccination, high-hospitalization counties such as Fulton County, GA (including Atlanta) and Jefferson County, AL (including Birmingham). At the other end, we observe high-vaccination, low-hospitalization counties such as Montgomery County, MD (including Rockville and Bethesda), Middlesex County, MA (including Cambridge), and King County, WA (including Seattle).

**Fig. 2.**
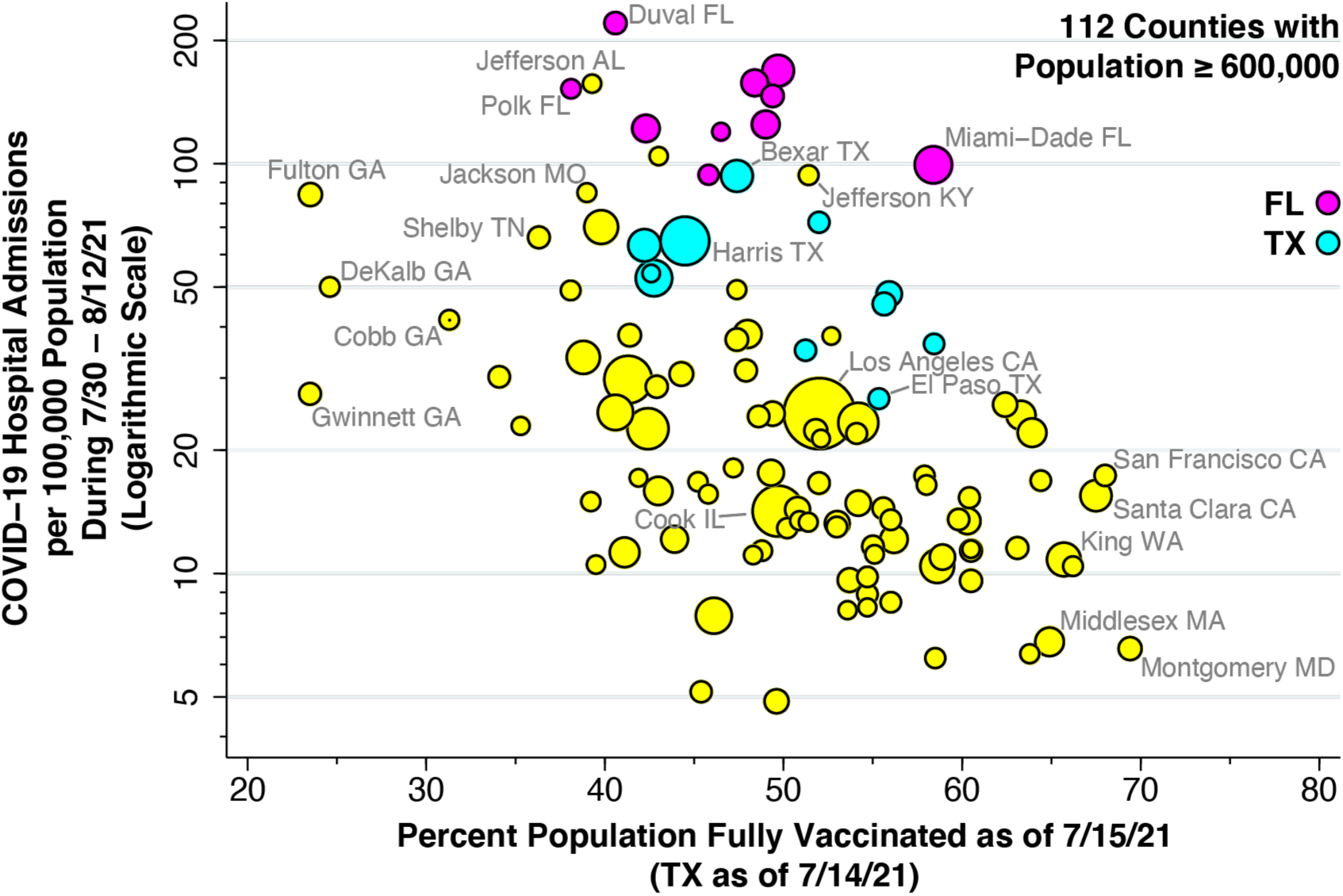
COVID-19 Hospital Admission Rate During 7/30 – 8/12/2021 Versus Vaccination Coverage as of 7/15/2021 in 112 U.S. Counties with Population ≥ 600,000. Hospital admission rate is measured on a logarithmic scale as admissions for confirmed cases of COVID-19 per 100,000 population. Vaccination coverage is measured as percent of population fully vaccinated. Vaccination coverage data for 11 Texas counties as of 7/14/2021. Florida counties highlighted in magenta. Texas counties highlighted in cyan. Size of data point proportional to county population.

Fig. 3 below plots hospital admissions per 100 cases in relation to vaccination coverage. While Miami-Dade County appeared to be an outlier in Figs. 1 and 2, with high incidence and hospitalization rates, Fig. 3 shows that its hospitalization-to-case ratio, an indicator of case severity, is in line with its 58.4 percent vaccination coverage.

**Fig. 3.**
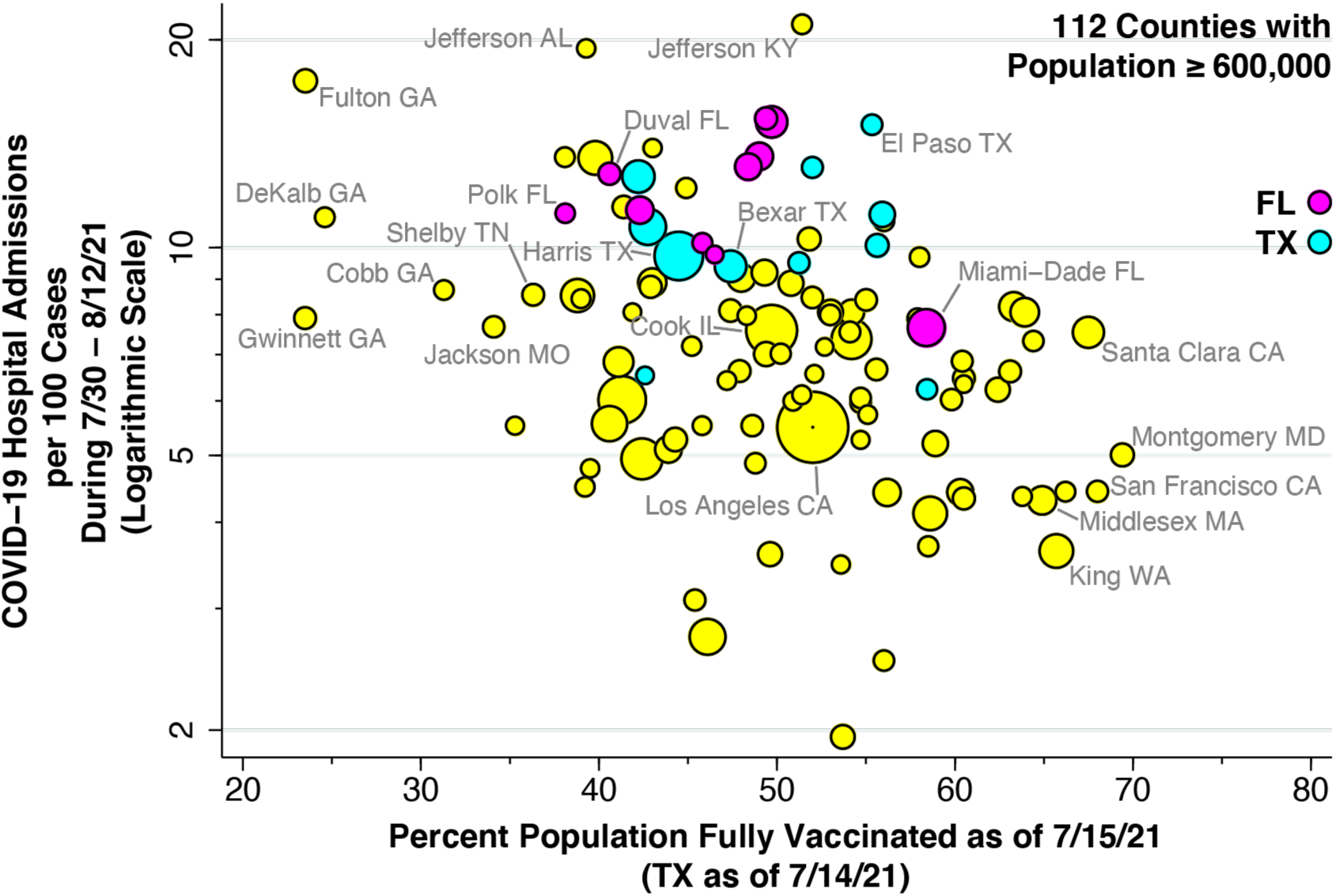
COVID-19 Hospital Admissions per 100 Cases During 7/30 – 8/12/2021 Versus Vaccination Coverage as of 7/15/2021 in 112 U.S. Counties with Population ≥ 600,000. COVID-19 hospital admissions per 100 cases is measured on a logarithmic scale as 100 times the ratio of the COVID-19 hospital admissions rate to the COVID-19 incidence rate. Vaccination coverage is measured as percent of population fully vaccinated. Vaccination coverage data for 11 Texas counties as of 7/14/2021. Florida counties highlighted in magenta. Texas counties highlighted in cyan. Size of data point proportional to county population.

Table 2 below provides our regression results for each of the three dependent variables. The estimated value of *β* = –0.0283 in the top panel means that a 10-percentage-point increase in vaccination coverage was associated with a 28.3% decrease in COVID-19 incidence (95% confidence interval, 16.8 – 39.7%). The estimated value of *β* = –0.0449 in the top panel means that a 10-percentage-point increase in vaccination coverage was associated with a 44.9% decrease in COVID-19 hospital admission rates (95% CI, 28.8 – 61.0%), while the estimated value of *β* = –0.0166 in the bottom panel means that the same 10-percentage-point increase in vaccination coverage was associate with a 16.6% decrease in COVID-19 hospitalizations per 100 cases (95% CI, 8.4 – 24.8%). The fact that the estimate of *β* displayed in the bottom panel equals the difference in the estimates of *β* derived from the other two panels is a direct consequence of our log-linear regression specification.

**Table 2.** Model Parameter Estimates for the Three Dependent Variables of Interest*.

|  |  |  |  |
| --- | --- | --- | --- |
| *COVID-19 Incidence* |  |  |  |
| Parameter | Model 1 | Model 2 | Model 3 |
| $\alpha$ | 7.224<br>(0.294) | 7.146<br>(0.305) | 6.819<br>(0.210) |
| $\beta$ | -0.0283<br>(0.0058) | -0.0254<br>(0.0060) | -0.0237<br>(0.0041) |
| $\mu_{FL}$ | | | 1.346<br>(0.134) |
| $\mu_{TX}$ | | | 0.588<br>(0.128) |
| $R^2$ | 0.179 | 0.139 | 0.600 |
| *COVID-19 Hospitalization Rate* |  |  |  |
| Parameter | Model 1 | Model 2 | Model 3 |
| $\alpha$ | 5.438<br>(0.413) | 5.314<br>(0.425) | 4.860<br>(0.290) |
| $\beta$ | -0.0449<br>(0.0081) | -0.0415<br>(0.0084) | -0.0386<br>(0.0056) |
| $\mu_{FL}$ | | | 1.861<br>(0.185) |
| $\mu_{TX}$ | | | 0.989<br>(0.176) |
| $R^2$ | 0.218 | 0.182 | 0.634 |
| *Hospital Admissions per 100 Cases* |  |  |  |
| Parameter | Model 1 | Model 2 | Model 3 |
| $\alpha$ | 2.821<br>(0.211) | 2.775<br>(0.218) | 2.647<br>(0.194) |
| $\beta$ | -0.0166<br>(0.0041) | -0.0161<br>(0.0043) | -0.0149<br>(0.0038) |
| $\mu_{FL}$ | | | 0.515<br>(0.124) |
| $\mu_{TX}$ | | | 0.401<br>(0.118) |
| $R^2$ | 0.128 | 0.114 | 0.298 |
| *Standard errors shown in parentheses below each parameter estimate. All estimates significantly different from 0 at the level $p = 0.001$ or lower. | | | |

The column corresponding to Model 2 in Table 2 shows insignificant changes in the estimated values of *β* when we ran a population-weighted regression rather than ordinary least squares. The results in the column corresponding to Model 3 demonstrate that the estimates of *β* remained significant even when we included the binary indicator variables for Florida and Texas.

### Ancillary Analyses

Table A3 in the Appendix displays the results of re-estimation of Models 1 through 3 on the alternative, expanded database of 138 counties with population ≥ 500,000. The results were virtually identical to those reported in Table 2 above.

Fig. 4 below displays the relation between the test positivity rate and vaccination coverage among the 112 counties in our principal sample. The scatterplot shows an inverse relation comparable to that shown in Figure 1.

**Fig. 4.**
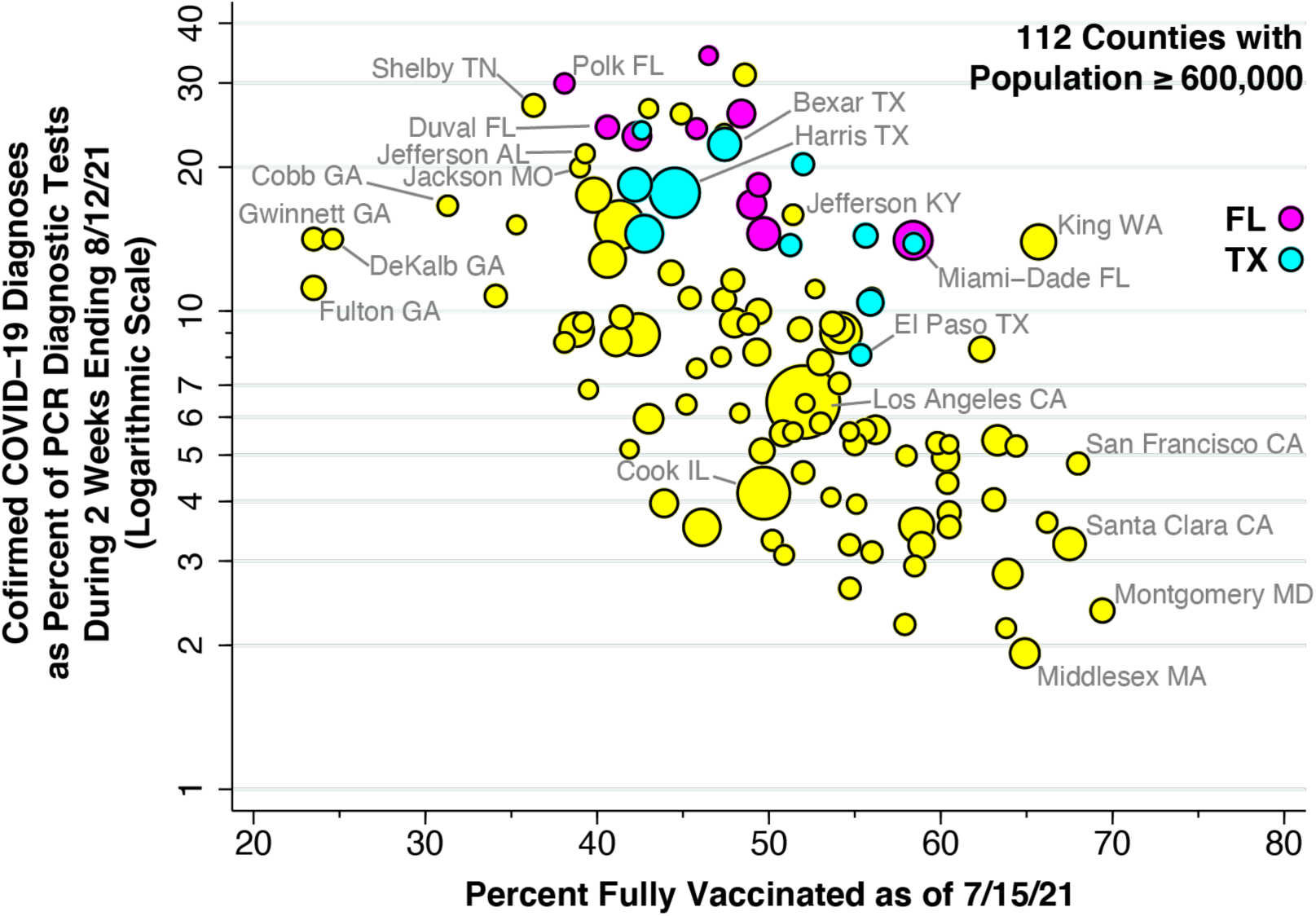
COVID-19 Test Positivity During 7/30 – 8/12/2021 Versus Vaccination Coverage as of 7/15/2021 in 112 U.S. Counties with Population ≥ 600,000. Test positivity is measured as 100% × the ratio of confirmed COVID-19 cases to total PCR diagnostic tests performed during 7/30 – 8/12/21. Vaccination coverage is measured as percent of population fully vaccinated. Vaccination coverage data for 11 Texas counties as of 7/14/2021. Florida counties highlighted in magenta. Texas counties highlighted in cyan. Size of data point proportional to county population.

Table 3 shows the estimates of the parameter *β* for the two alternative dependent variables: the test positivity rate; and the death rate. The estimates relating the test positivity to the vaccine participation rate were larger in absolute value than those where COVID-19 incidence as the dependent variable, as shown in Table 2. In particular, the OLS regression of the logarithm of the test positivity rate (Model 1) had a slope *β* = –0.0441, with 95% confidence interval (CI) [–0.0556, –0.0325], whereas the corresponding OLS regression of the logarithm of the COVID-19 incidence rate had a slope *β* = –0.0283, with 95% CI [–0.0397, –0.0168]. The difference between the two estimates of *β* approached statistical significance (two-sided Z-test, p = 0.054).

**Table 3.** Estimates of the Parameter *β* for Two Alternative Dependent Variables*.

| **Table 3. Estimates of the Parameter $\beta$ for Two Alternative Dependent Variables\*** |  |  |
| --- | --- | --- |
| **Model** | **Test Positivity Rate¶** | **Death Rate§** |
| 1 | −0.0441<br>(0.0058) | −0.0464<br>(0.0089) |
| 2 | −0.0423<br>(0.0060) | −0.0426<br>(0.0092) |
| 3 | −0.0407<br>(0.0047) | −0.0386<br>(0.0055) |
| *Standard errors are shown below each estimate of the parameter $\beta$, relating the logarithm of the dependent variable $Y$ to vaccination coverage $X$.<br>¶Measured as the ratio of the number of COVID-19 cases to the number of PCR diagnostic tests performed during the 2-week period ending 8/12/21, expressed as a percentage.<br>§Measured as the number of COVID-19 deaths per 100,000 population during the 4-week period from 8/20 through 9/14/21. |  |  |

As further indicated in Table 3, the estimates of the parameter *β* relating the death rate to the vaccine participation rate were all different from zero at the significance level p < 0.001. The estimate of *β* in Model 3, where binary indicators for Florida and Texas were included, was somewhat lower, principally because Florida counties had a more than double the COVID-19 death rate (*μ*_*FL*_ = 2.32, with 95% CI [1.96, 2.68]).

Figure 5 shows estimates of the parameter *β* under Model 4 (log *Y* = *α* + *β X* + *γZ*), where the dependent variable *Y* was the logarithm of the COVID-19 hospital admission rate, and where the additional covariate *Z* was one of six independent variables. The datapoint at the extreme left corresponds to the base case (Model 1) where the covariate *Z* was omitted. In all cases, the estimate of *β* was significantly different from zero at the level p < 0.001. While the point estimate of *β* was somewhat lower in absolute value when the social vulnerability index or diabetes prevalence were included as covariates, none of the estimates of *β* were significantly different from the base-case estimate with no covariate. For diabetes prevalence, a two-sided Z-test of the difference in *β*’s gave p = 0.462.

**Fig. 5.**
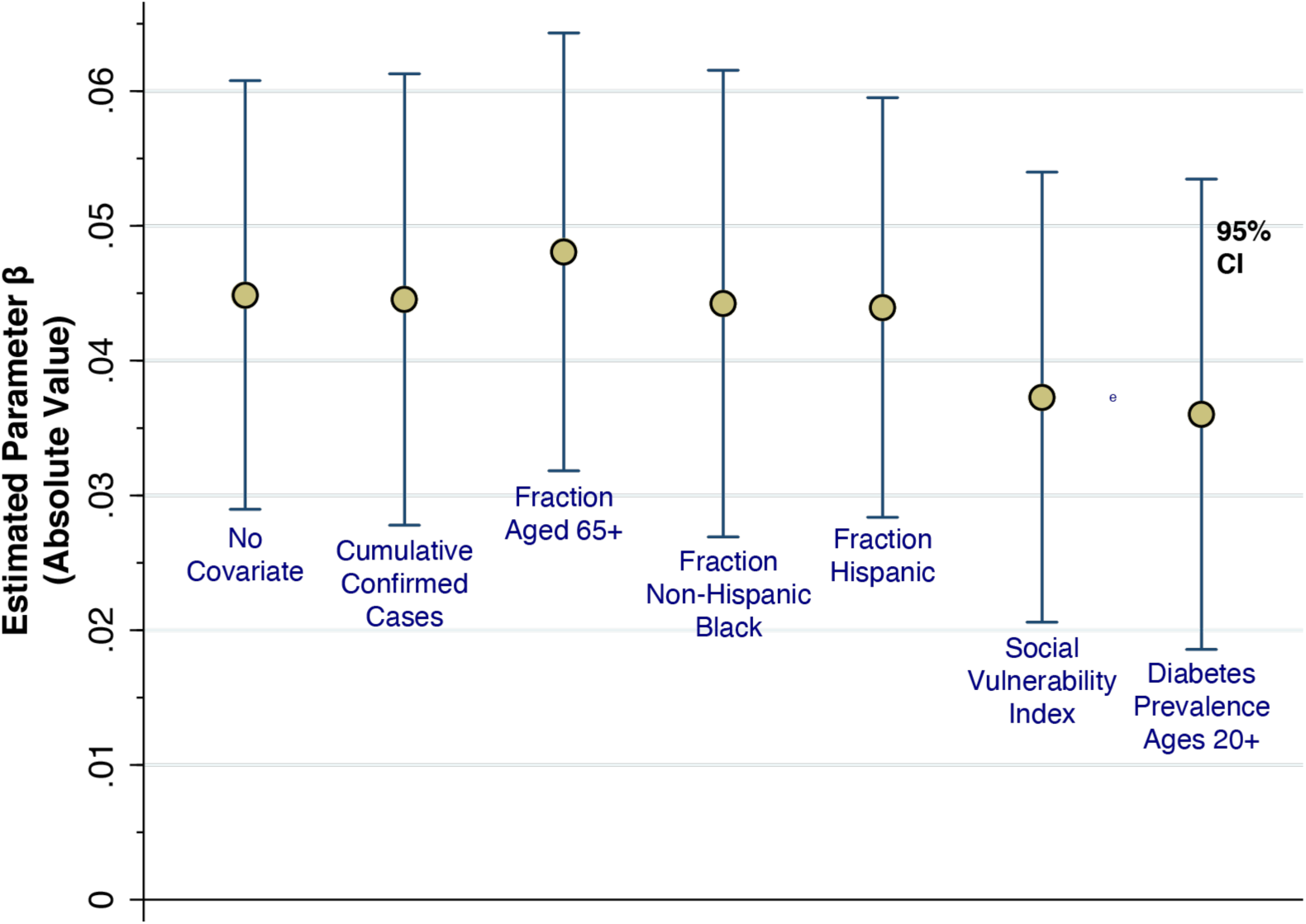
Effect of Including Covariates *Z* on the Estimated Coefficients *β* in Models of the Hospitalization Rate. We repeatedly estimated Model 4 (log *Y* = *α* + *β X* + *γZ*), where *Y* denotes COVID-19 hospital rate per 100,000 population in each county, *X* denotes the corresponding vaccination coverage, and *Z* denotes a county-specific covariate. Each datapoint corresponds to the absolute value of the estimated parameter *β*. The error bars indicate the 95% confidence intervals. The datapoint at the extreme left corresponds to the base case (Model 1) where the covariate *Z* was omitted.

Table A4 in the Appendix shows the corresponding estimates of the parameter *γ* under Model 4, where both the dependent variable *Y* and the additional covariate *Z* were varied. The cumulative number of COVID-19 cases per 100 persons through 6/30/21 had no significant effect on any of the dependent variables. What’s more, the number of cumulative confirmed cases per 100 population in the 10 Florida counties was indistinguishable from that of the remaining counties (t-test of group means with unequal variance, p = 0.49). The fraction Hispanic had a significant positive effect on both COVID-19 incidence and the hospitalization rate. The SVI score and diabetes prevalence had significant positive effects on both the hospitalization rate and the hospitalizations per 100 cases.

## Discussion

Numerous factors could have contributed to the substantial scatter of the datapoints seen in Figs. 1 through 3. In our ancillary analyses, we attempted to control for county-specific differences in demographic characteristics, as well as the prevalence of diabetes, a strong predictor of COVID-19 case severity.^22^ Apart from these factors, it is possible that differences in public policies, including prohibition of mandates on vaccination and mask-wearing in schools and workplaces in certain states, may have been contributory.^23^ A critical limitation of the current study is that the *COVID-19 Community Profile Report*, maintained at *healthdata*.*gov*,^12^ does not provide a detailed breakdown of our county-specific data on vaccination coverage, COVID-19 incidence and hospitalization rates by age group. Still, the persistence of clearly detectable differences between low- and high-vaccination counties – even with the low *R*^2^ statistics seen in the regression results in Table 2 – points to an important, identifiable deterrent effect of vaccinations on disease spread during the Delta surge.

Our analysis focused on the most populous counties in the U.S., comprising 44.4% of the total population. We excluded less populous, rural counties, where transmission dynamics are likely to be quite different,^24^ and where smaller population denominators tend to result in higher sampling variability. We thus avoided the pitfall of drawing biased conclusions from the study of rural and urban counties combined.^25^ While our choice of a population cutoff of 600,000 inhabitants is necessarily arbitrary, our principal results remained unchanged when we expanded our database by lowering the cutoff to 500,000 (Appendix Table A3).

While there is evidence that as many as one-third of COVID-19 suffers have no detectable antibodies against SARS-CoV-2,^26^ it is likely that those who experienced a sufficiently high viral load during their illness have acquired some degree of natural immunity. In that case, we would expect to observe a protective effect of higher rates of past infection on COVID-19 incidence and hospitalizations, even taking vaccination coverage into account. Yet the cumulative prevalence of confirmed COVID-19 infection had no significant effect on any of our principal dependent variables (Appendix Table A4). Nor did its inclusion in our regression Model 4 attenuate the effect of vaccination coverage (Fig. 5).

Counts of confirmed cases based on voluntary testing of symptomatic individuals are known to have significantly understated actual numbers of SARS-CoV-2 infections ^27 28^ This consideration at least raises the possibility that the extent of ascertainment of COVID-19 infections could be inversely correlated with a county’s vaccination coverage. The respective parameter estimates of *β* in Model 1 (log *Y* = *α* + *βX*) were –0.0283 when the dependent variable *Y* was the COVID-19 incidence rate (Table 2) and –0.0441 when the dependent variable was the test positivity rate (Table 3). The fact that the former estimate of *β* is algebraically greater than the latter implies that counties with higher vaccination coverage have performed *more* testing per capita. To maintain that an ascertainment bias is a valid explanation for the significant inverse relation seen in Fig. 1, one would have to posit that counties with higher vaccination coverage have been more aggressive in testing uninfected individuals while somehow detecting fewer infected individuals.

Our finding that COVID-19 death rates are inversely related to vaccination coverage needs is consistent with our results on hospitalization rates. Still, there is a substantial, highly variable delay between initial diagnosis and death, with the mean lag time for the original coronavirus on the order of 16 days.^29 30^ While the Delta variant appears to have a shorter incubation time from infection to symptoms,^31^ the time from symptoms to death is less well characterized. We measured COVID-19 incidence and hospitalization during 7/30 – 8/12/21, an observation interval starting two weeks after the mid-July cutoff date for ascertaining vaccination coverage. To accommodate the variable delay in mortality, we measured subsequent deaths during 8/20 – 9/16/21. However, we cannot be confident of a one-to-one mapping between cases diagnosed during 7/30 – 8/12/21 and deaths that occurred during 8/20 – 9/16/21.

Our scatterplots (Figs. 1 and 2) and regression results (Table 2, Model 3) suggest that the 10 Florida counties may be outliers, with rates of COVID-19 infections and hospitalizations significantly above the level expected for their observed vaccination coverage. Our finding that cumulative infections through 6/30/21 did not predict subsequent COVID-19 incidence (Appendix Table A4) fails to support the hypothesis that Florida’s high rates of infection and hospitalization have been the result of a lower level of pre-existing, acquired population immunity. Further research on the impact of Florida’s statewide policies is needed.

The log-linear specification of our regression models implies that incremental increases in vaccination coverage have the strongest protective effect at low baseline rates of vaccination coverage. Given the parameter estimates of *α* = 0.7724 and *β* = 0.0283 in Model 1 (as shown in Table 2), an increase in vaccination coverage from 10 to 20 percent of the population would reduce the 14-day COVID-19 incidence by 255 per 100,000. By contrast, an increase in coverage from 50 to 60 percent would reduce 14-day incidence by 82 per 100,000. This built-in nonlinearity is consistent with the predictions of a variety of compartmental models of infectious disease propagation.^32^ Still, our non-parametric descriptive analysis comparing the lower and upper halves of the vaccination coverage distribution makes clear that our results do not depend on the specification of a particular parametric model.

Our results do not bear directly on the existence or extent of breakthrough infections among vaccinated individuals, or on the capacity of such individuals to transmit their infections to others. Nor do they shed light on the question of waning vaccine effectiveness. They do suggest, however, that these phenomena are not sufficiently important on a large scale to completely attenuate the inverse relationship between COVID-19 incidence and vaccination coverage seen here. Our findings add large-scale, population-level evidence to the growing body of individual-level studies concluding that vaccination remains highly effective in preventing severe disease.^33 34^

In view of the persistence of a critical mass of unvaccinated individuals throughout the United States, complete elimination of COVID-19 in the foreseeable future is simply not in the offing. The more realistic short-term goal is to reduce disease severity. Our quantitative findings indicate that even a marginal increase in vaccination coverage would substantially reduce the proportion of infected individuals who end up in the hospital. That, in turn, would markedly reduce the strain on the country’s healthcare resources that was seen during the Delta surge.^35-37^

## Data Availability

The sources of data for this study are publicly accessible via the Internet at the URLs cited in the reference section. We have posted our data analyses at the Open Science Framework (OSF) in a project entitled 112-County COVID-19 Incidence-Vaccination Study (https://osf.io/wtb6j/).

## Acknowledgments

This article represents the sole opinion of its author and does not necessarily represent the opinions of the Massachusetts Institute of Technology, Eisner Health, or any other organization or individual. The comments of Jason L. Salemi on an earlier version of this article^38^ are gratefully acknowledged.

## Human Subjects Declaration

This study relies exclusively on anonymized, publicly available data that contain no individual identifiers.

## Data Availability Statement

The sources of data for this study are publicly accessible via the Internet links cited in the reference section. We have posted our data analyses at the Open Science Framework (OSF) in a project entitled *112-County COVID-19 Incidence-Vaccination Study* (https://osf.io/wtb6j/).

## Appendix

**Fig. A1.**
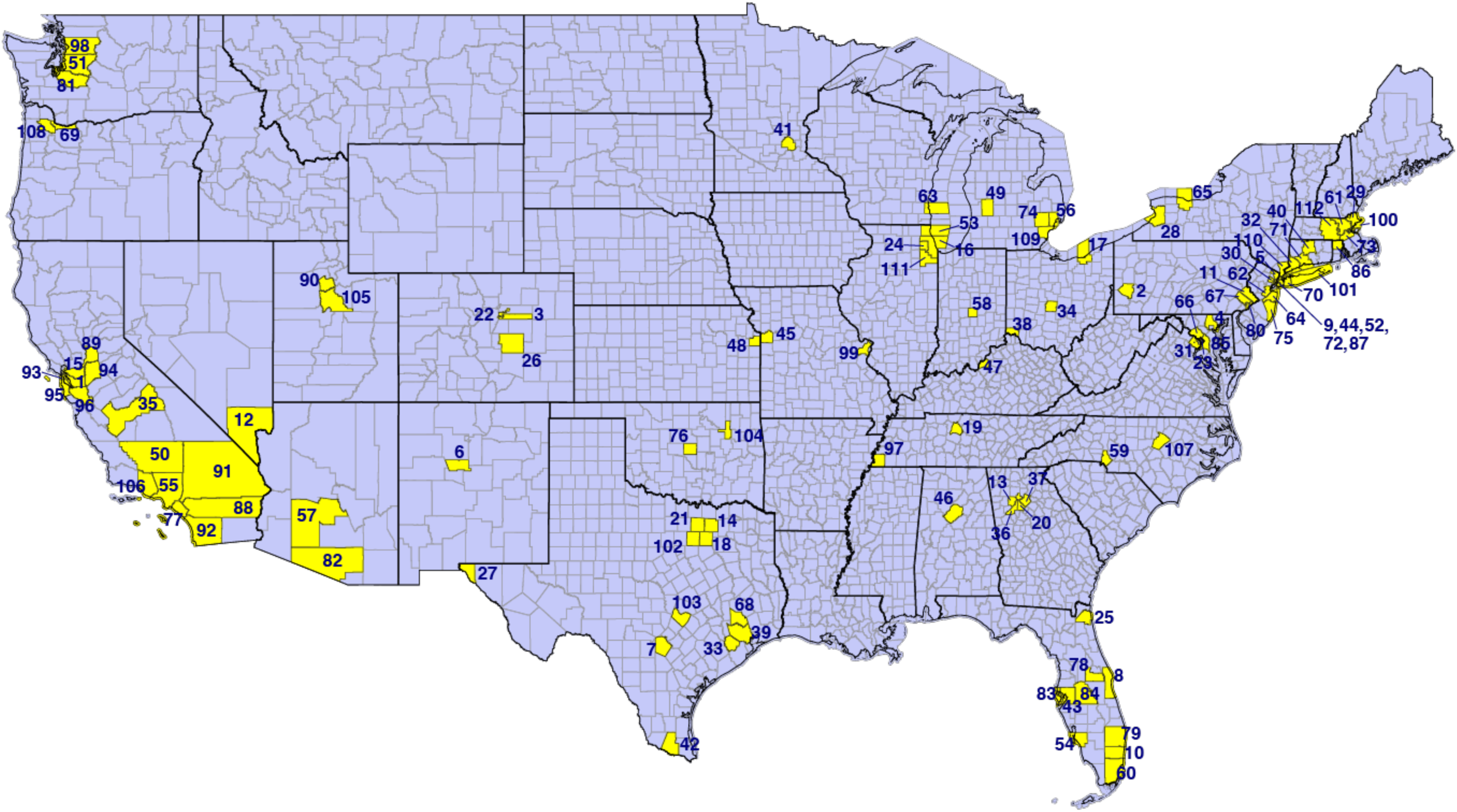
112 U.S. Counties with Population ≥ 600,000. All counties are numbered in accordance with the legend below. State boundaries are indicated by the thicker black lines.

**Legend to Fig. A1.**
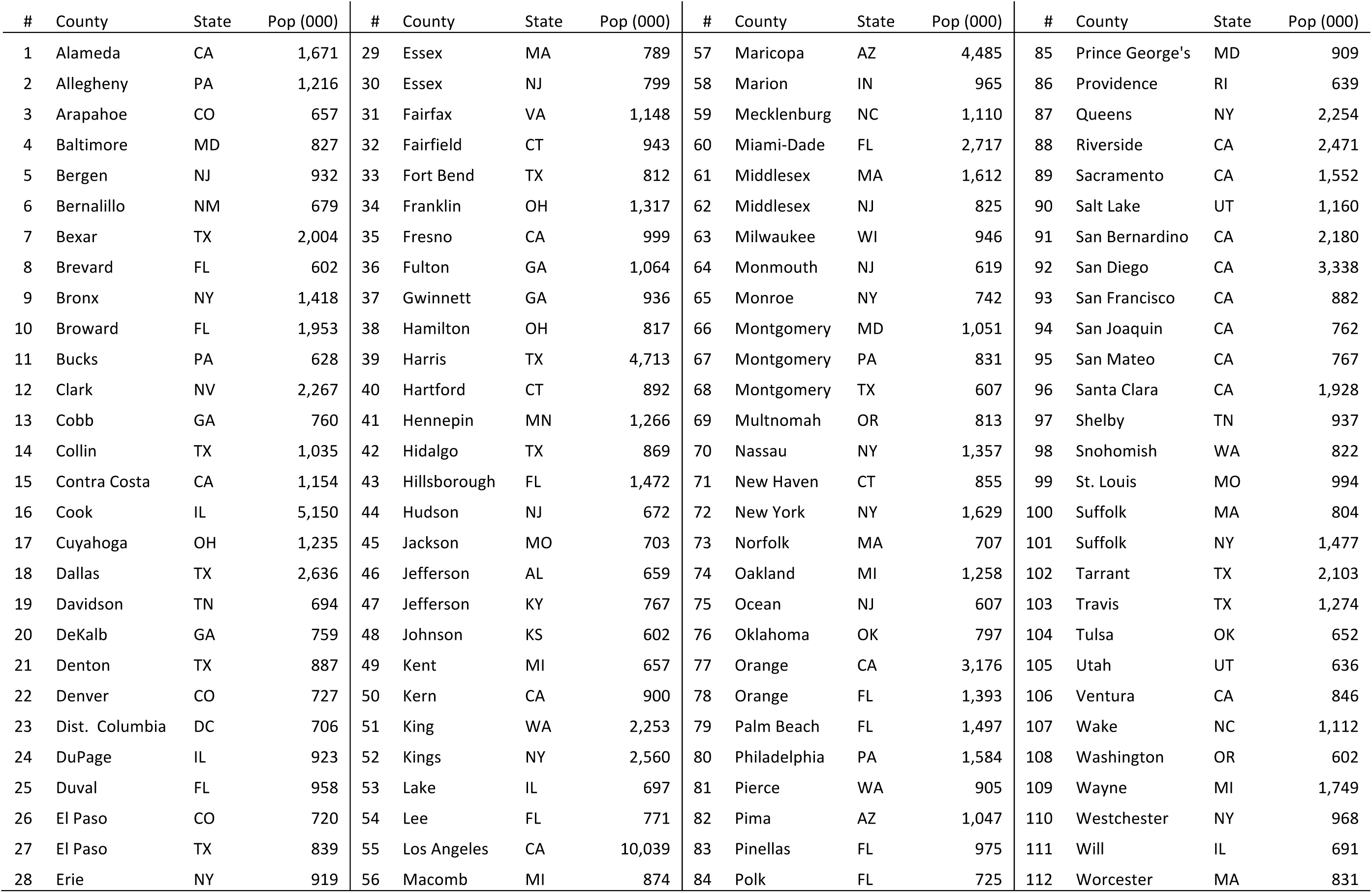
Counties, along with their population in thousands, are listed in alphabetical order.

**Table A1.**
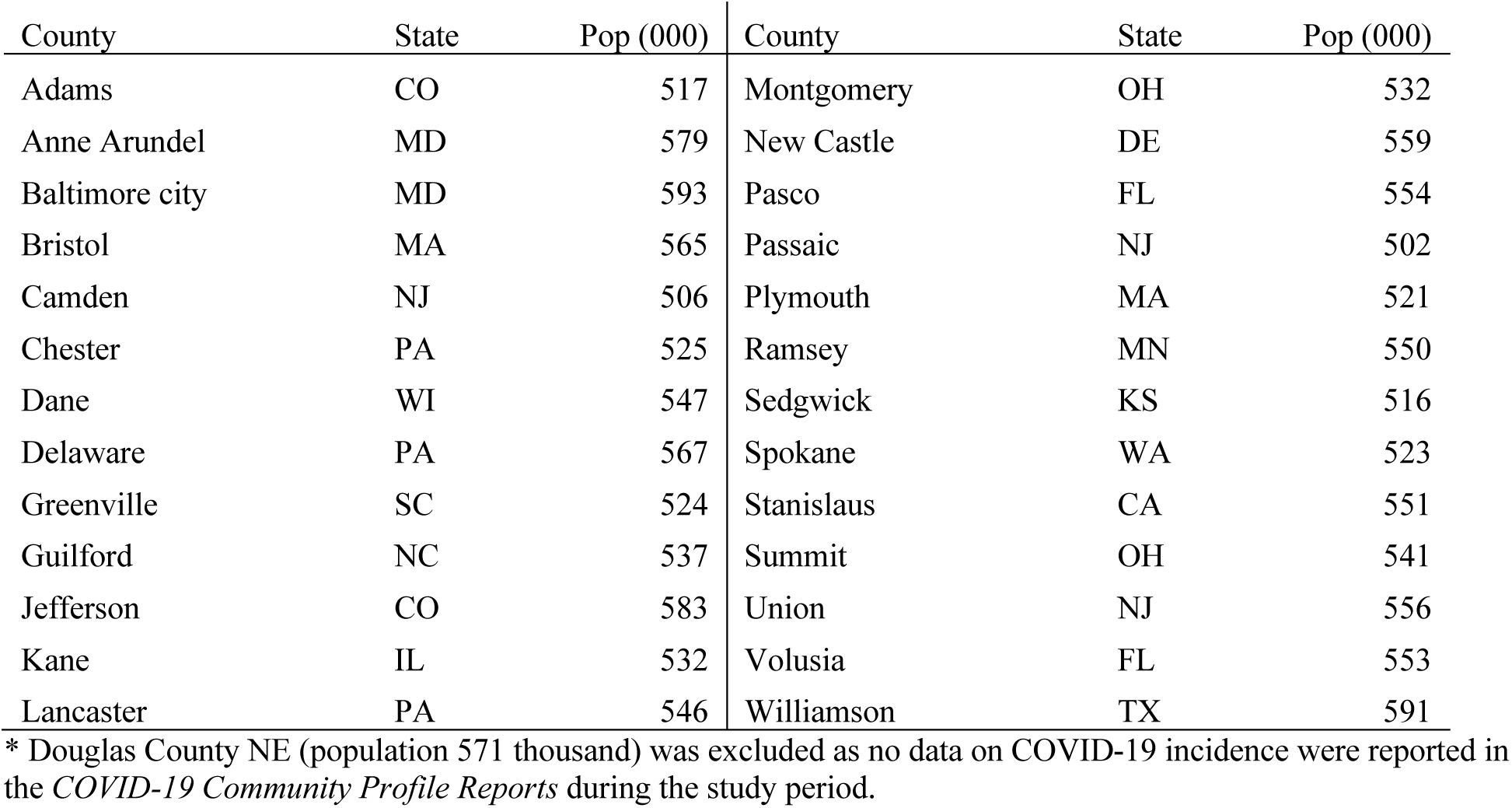
26 Additional Counties Included in Ancillary Analysis*.

**Table A2.** Summary Statistics of Variables Used in the Principal and Ancillary Analyses.

| Variable | No. Obs. | Frequency | Mean | Std. Dev. |
| --- | --- | --- | --- | --- |
| *Vaccination Coverage* (% Fully Vaccinated as of 7/15/21) | 112 |  | 49.99 | 9.33 |
| *COVID-19 Incidence* (Cases per 100,000 Population During 2 Weeks Ending 8/12/21) | 112 |  | 412.22 | 307.67 |
| *COVID-19 Hospitalization Rate* (Admissions per 100,000 Population During 2 Weeks Ending 8/12/21) | 112 |  | 37.93 | 41.71 |
| *Hospitalizations per 100 Cases* ($100 \times \text{Incidence} / \text{Hospitalization Rate}$) | 112 | | 8.01 | 3.51 |
| *Florida County* (Binary Indicator Variable) | 112 | 10 |  |  |
| *Texas County* (Binary Indicator Variable) | 112 | 11 |  |  |
| *Test Positivity Rate* (Confirmed Cases per 100 PCR Tests Performed During 2 Weeks Ending 8/12/21) | 112 |  | 10.63 | 7.42 |
| *Death Rate* (COVID-19 Deaths per 100,000 Population During 8/20 – 9/14/21) | 112 |  | 11.34 | 16.46 |
| *Cumulative Cases* (Confirmed COVID-19 Cases per 100 Population as of 6/30/2021) | 112 |  | 10.00 | 2.53 |
| *Fraction 65+* (Fraction of Population Aged 65 or Older) | 112 |  | 0.1513 | 0.0326 |
| *Fraction Non-Hispanic Black* (Fraction of Population Reported as Non-Hispanic Black) | 112 |  | 0.1519 | 0.1253 |
| *Fraction Hispanic* (Fraction of Population Reported as Hispanic) | 112 |  | 0.2230 | 0.1674 |
| *SVI Score* (CDC's Social Vulnerability Index, Range 0–1) | 112 |  | 0.5180 | 0.2345 |
| *Diabetes Prevalence* (Prevalence of Diabetes per 100 Persons Aged 20+, 2018) | 106 |  | 8.91 | 1.49 |

**Table A3.** Model Parameter Estimates for the Three Dependent Variables of Interest: Alternative Database of 138 Counties with Population Exceeding 500,000*§.

|  |  |  |  |
| --- | --- | --- | --- |
| *COVID-19 Incidence* |  |  |  |
| Parameter | Model 1 | Model 2 | Model 3 |
| $\alpha$ | 7.217<br>(0.275) | 7.144<br>(0.281) | 6.795<br>(0.194) |
| $\beta$ | -0.0291<br>(0.0054) | -0.0259<br>(0.0056) | -0.0242<br>(0.0038) |
| $\mu_{FL}$ | | | 1.378<br>(0.122) |
| $\mu_{TX}$ | | | 0.687<br>(0.121) |
| $R^2$ | 0.173 | 0.137 | 0.610 |
| *COVID-19 Hospitalization Rate* |  |  |  |
| Parameter | Model 1 | Model 2 | Model 3 |
| $\alpha$ | 5.430<br>(0.378) | 5.317<br>(0.386) | 4.849<br>(0.261) |
| $\beta$ | -0.0459<br>(0.0075) | -0.0422<br>(0.0076) | -0.0393<br>(0.0051) |
| $\mu_{FL}$ | | | 1.870<br>(0.164) |
| $\mu_{TX}$ | | | 1.063<br>(0.163) |
| $R^2$ | 0.217 | 0.183 | 0.644 |
| *Hospital Admissions per 100 Cases* |  |  |  |
| Parameter | Model 1 | Model 2 | Model 3 |
| $\alpha$ | 2.818<br>(0.191) | 2.778<br>(0.196) | 2.660<br>(0.177) |
| $\beta$ | -0.0168<br>(0.0038) | -0.0161<br>(0.0043) | -0.0151<br>(0.0035) |
| $\mu_{FL}$ | | | 0.493<br>(0.111) |
| $\mu_{TX}$ | | | 0.376<br>(0.110) |
| $R^2$ | 0.128 | 0.115 | 0.281 |
| *Standard errors shown in parentheses below each parameter estimate. All estimates significantly different from 0 at the level $p = 0.001$ or lower.<br>§Excludes Douglas County NE, population 571,000, for which no data on COVID-19 incidence were reported. | | | |

Note that cum cases had insignificant gamma. Make table of gamma values for all Z’s for three models. Table has 3 columns for three dependent variables and 6 rows for each Z.

**Table A4.**
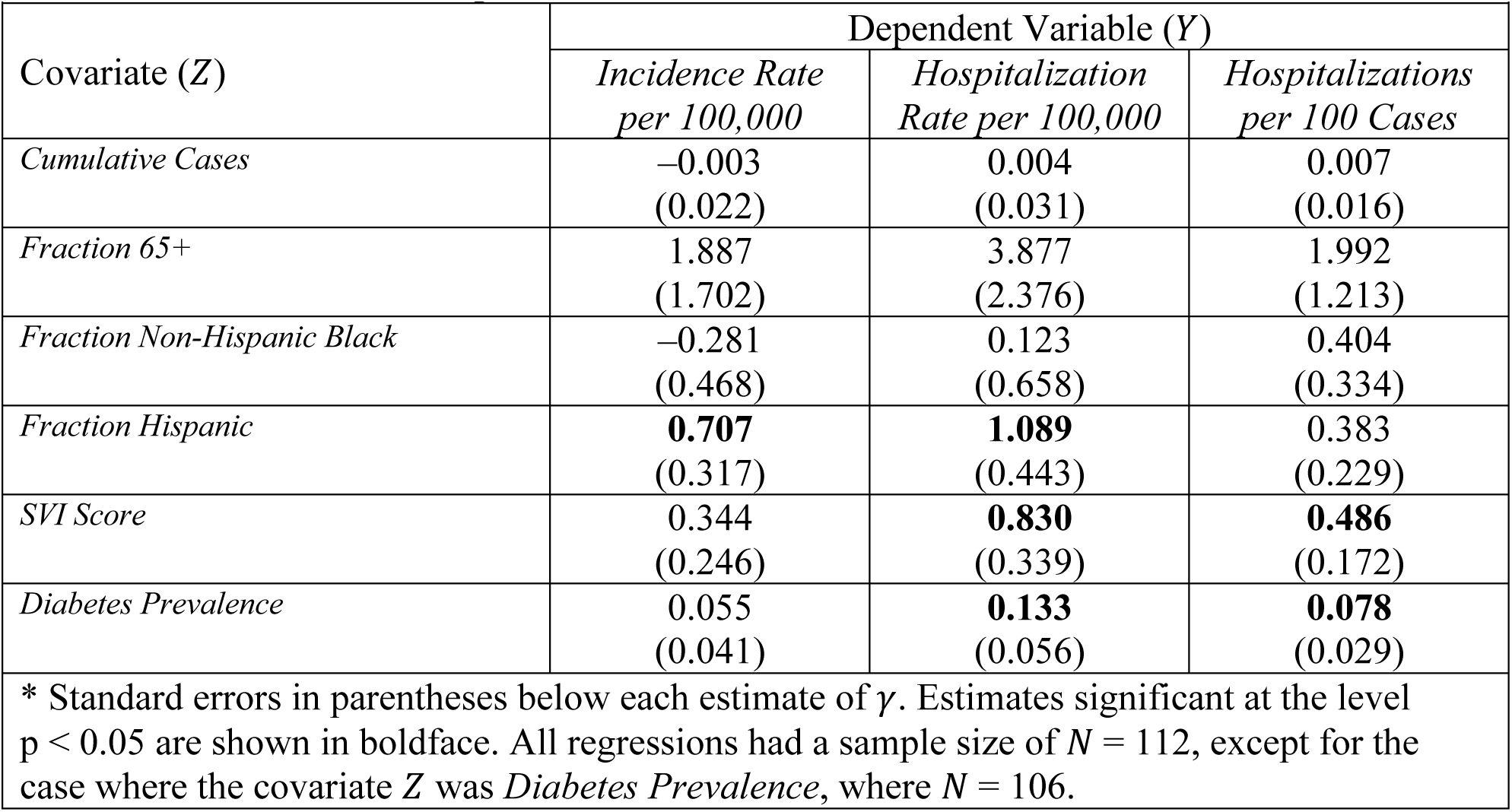
Estimates of the Parameter *γ* for Model 4 (log *Y* = *α* + *βX* + *γZ*) for Each Dependent Variable *Y* and Covariate *Z*\*.

| **Table A4. Estimates of the Parameter $\gamma$ for Model 4 ($\log Y = \alpha + \beta X + \gamma Z$) for Each Dependent Variable $Y$ and Covariate $Z$\*** |  |  |  |
| --- | --- | --- | --- |
| Covariate ($Z$) | Dependent Variable ($Y$) | | |
|  | *Incidence Rate*<br>*per 100,000* | *Hospitalization*<br>*Rate per 100,000* | *Hospitalizations*<br>*per 100 Cases* |
| *Cumulative Cases* | −0.003<br>(0.022) | 0.004<br>(0.031) | 0.007<br>(0.016) |
| *Fraction 65+* | 1.887<br>(1.702) | 3.877<br>(2.376) | 1.992<br>(1.213) |
| *Fraction Non-Hispanic Black* | −0.281<br>(0.468) | 0.123<br>(0.658) | 0.404<br>(0.334) |
| *Fraction Hispanic* | **0.707**<br>(0.317) | **1.089**<br>(0.443) | 0.383<br>(0.229) |
| *SVI Score* | 0.344<br>(0.246) | **0.830**<br>(0.339) | **0.486**<br>(0.172) |
| *Diabetes Prevalence* | 0.055<br>(0.041) | **0.133**<br>(0.056) | **0.078**<br>(0.029) |
| * Standard errors in parentheses below each estimate of $\gamma$. Estimates significant at the level $p < 0.05$ are shown in boldface. All regressions had a sample size of $N = 112$, except for the case where the covariate $Z$ was *Diabetes Prevalence*, where $N = 106$. | | | |

## References

1. GISAID. Tracking of Variants https://www.gisaid.org/hcov19-variants/: Accessed July 9 2021.

2. Stein R, Wroth C, Fast A. Where Are The Newest COVID Hot Spots? Mostly Places With Low Vaccination Rates. https://www.npr.org/sections/health-shots/2021/07/09/1014512213/covid-is-surging-in-new-hotspots-driven-by-low-vaccination-rates: NPR, July 9 2021.

3. Rattner N. Here’s a map showing where low vaccination rates meet high case counts as U.S. Covid infections surge. https://www.cnbc.com/2021/07/13/covid-cases-rise-in-us-counties-with-low-vaccination-rates-as-delta-variant-spreads.html: CNBC, July 13 2021.

4. Leatherby L. As Covid Cases Rise All Over U.S., Lower Vaccination Rates Point to Worse Outcomes. https://www.nytimes.com/interactive/2021/07/31/us/covid-delta-cases-deaths.html: New York Times, July 31 2021.

5. Nirappil F, Keating D, Aguilar M, et al. Spread of delta variant ignites covid hot spots in highly vaccinated parts of the U.S., Post analysis finds. https://www.washingtonpost.com/health/interactive/2021/vaccinated-counties-delta-hotspots/: Washington Post, August 12 2021.

6. Lange B, Gerigk M, Tenenbaum T. Breakthrough Infections in BNT162b2-Vaccinated Health Care Workers. N Engl J Med 2021 doi: 10.1056/NEJMc2108076 [published Online First: 2021/08/19]

7. Nanduri S, Pilishvili T, Derado G, et al. Effectiveness of Pfizer-BioNTech and Moderna Vaccines in Preventing SARS-CoV-2 Infection Among Nursing Home Residents Before and During Widespread Circulation of the SARS-CoV-2 B.1.617.2 (Delta) Variant - National Healthcare Safety Network, March 1-August 1, 2021. MMWR Morb Mortal Wkly Rep 2021;70(34):1163–66. doi: 10.15585/mmwr.mm7034e3 [published Online First: 2021/08/27]

8. Israel A, Merzon E, Schaeffer AA, et al. Elapsed time since BNT162b2 vaccine and risk of SARS-CoV-2 infection in a large cohort. https://www.medrxiv.org/content/10.1101/2021.08.03.21261496v1: medRxiv, August 5 2021.

9. Tapp T. In Los Angeles, Breakthrough Infections Are Now 30% Of All New Covid Cases Amid Delta Surge. https://deadline.com/2021/08/los-angeles-breakthrough-infections-covid-amount-cases-1234818477/: Deadline, August 19 2021.

10. Brown CM, Vostok J, Johnson H, et al. Outbreak of SARS-CoV-2 Infections, Including COVID-19 Vaccine Breakthrough Infections, Associated with Large Public Gatherings — Barnstable County, Massachusetts, July 2021. MMWR 2021;70(Early Release):ePub July 30. doi: 10.15585/mmwr.mm7031e2

11. Shamier MC, Tostmann A, Bogers S, et al. Virological characteristics of SARS-CoV-2 vaccine breakthrough infections in health care workers. https://www.medrxiv.org/content/10.1101/2021.08.20.21262158v1: medRxiv, August 21 2021.

12. healthdata.gov. COVID-19 Community Profile Report. https://healthdata.gov/Health/COVID-19-Community-Profile-Report/gqxm-d9w9: U.S. Department of Health and Human Resources, last accessed 8/17/2021 2021.

13. Democrat and Chronicle. Texas COVID-19 Vaccine Tracker. https://web.archive.org/web/20210714234135/https://data.democratandchronicle.com/covid-19-vaccine-tracker/texas/48/: Last accessed August 16 2021.

14. Centers for Disease Control and Prevention. CDC Social Vulnerability Index (SVI). https://data.cdc.gov/Health-Statistics/CDC-Social-Vulnerability-Index-SVI-/u6k2-rtt3: Data.cdc.gof 2020.

15. New York Times. covid-19-data. https://github.com/nytimes/covid-19-data: Accessed August 26 2021.

16. New York Department of Health and Mental Hygiene. nyc health/coronavirus-data. https://github.com/nychealth/coronavirus-data/tree/master/: Accessed August 28 2021.

17. Centers for Disease Control and Prevention. Diabetes Atlas. https://gis.cdc.gov/grasp/diabetes/DiabetesAtlas.html2020.

18. Montgomery B. Florida leads U.S. in COVID cases amid Delta-driven surge. https://www.axios.com/local/tampa-bay/2021/07/19/florida-covid-cases-delta-variant: Axios Tampa Bay, mJuly 19 2021.

19. ABC News. COVID-19 cases, hospitalizations surge in Florida as delta variant spreads (Video). https://abcnews.go.com/Health/video/covid-19-cases-hospitalizations-surge-florida-delta-variant-79016389: mJuly 21 2021.

20. Lopez A. Delta Variant Is Making Up More Than 75% Of New COVID-19 Cases In Texas, Health Officials Say. https://www.kut.org/covid-19/2021-08-04/delta-variant-is-making-up-more-than-75-of-new-covid-19-cases-in-texas-health-officials-say: KUT 90.5, August 4 2021.

21. Lin R-GI, Money L. California doing much better with Delta variant than Florida, Texas. Here’s why. https://www.latimes.com/california/story/2021-08-11/despite-surge-california-doing-much-better-with-delta-variant-than-florida-texas-heres-why: Los Angeles Times, August 11 2021.

22. Gregory JM, Slaughter JC, Duffus SH, et al. COVID-19 Severity Is Tripled in the Diabetes Community: A Prospective Analysis of the Pandemic’s Impact in Type 1 and Type 2 Diabetes. Diabetes Care 2021;44(2):526–32. doi: 10.2337/dc20-2260 [published Online First: 2020/12/04]

23. Mulligan K, Harris JE. COVID-19 Vaccination Mandates for School and Work Are Sound Public Policy. https://healthpolicy.usc.edu/research/covid-19-vaccination-mandates-for-school-and-work-are-sound-public-policy/: University of Southern California, Leanard D. Schaeffer Center for Health Policy & Economics, July 7 2021.

24. Li Y, Hu T, Gai X, et al. Transmission Dynamics, Heterogeneity and Controllability of SARS-CoV-2: A Rural-Urban Comparison. Int J Environ Res Public Health 2021;18(10) doi: 10.3390/ijerph18105221 [published Online First: 2021/06/03]

25. Subramanian SV, Kumar A. Increases in COVID-19 are unrelated to levels of vaccination across 68 countries and 2947 counties in the United States. Eur J Epidemiol 2021 doi: 10.1007/s10654-021-00808-7 [published Online First: 2021/10/01]

26. Liu W, Russell RM, Bibollet-Ruche F, et al. Predictors of Nonseroconversion after SARS-CoV-2 Infection. Emerg Infect Dis 2021;27(9):2454–58. doi: 10.3201/eid2709.211042 [published Online First: 2021/07/02]

27. Oran DP, Topol EJ. Prevalence of Asymptomatic SARS-CoV-2 Infection : A Narrative Review. Ann Intern Med 2020;173(5):362–67. doi: 10.7326/M20-3012 [published Online First: 2020/06/04]

28. Havers FP, Reed C, Lim TW, et al. Seroprevalence of Antibodies to SARS-CoV-2 in Six Sites in the United States, March 23-May 3, 2020. https://www.medrxiv.org/content/10.1101/2020.06.25.20140384v1: MedRxiv June 26, 2020 2020.

29. Harris JE. Reopening Under COVID-19: What to Watch For. http://web.mit.edu/jeffrey/harris/HarrisJE_WP3_COVID19_WWF_6-May-2020.pdf: May 12, 2020 2020.

30. Harris JE. COVID-19 Case Mortality Rates Continue to Decline in Florida. https://www.medrxiv.org/content/10.1101/2020.08.03.20167338v1: medRxiv August 4, 2020 2020.

31. Kang M, Xin H, Ali ST, et al. Transmission dynamics and epidemiological characteristics of Delta variant infections in China. https://www.medrxiv.org/content/10.1101/2021.08.12.21261991v1: medRxiv, August 13 2021.

32. Williams JR, Nokes DJ, Medley GF, et al. The transmission dynamics of hepatitis B in the UK: a mathematical model for evaluating costs and effectiveness of immunization programmes. Epidemiol Infect 1996;116(1):71–89. doi: 10.1017/s0950268800058970 [published Online First: 1996/02/01]

33. Rosenberg ES, Holtgrave DR, Dorabawila V, et al. New COVID-19 Cases and Hospitalizations Among Adults, by Vaccination Status - New York, May 3-July 25, 2021. MMWR Morb Mortal Wkly Rep 2021;70(34):1150–55. doi: 10.15585/mmwr.mm7034e1 [published Online First: 2021/08/27]

34. Tenforde MW, Self WH, Naioti EA, et al. Sustained Effectiveness of Pfizer-BioNTech and Moderna Vaccines Against COVID-19 Associated Hospitalizations Among Adults - United States, March-July 2021. MMWR Morb Mortal Wkly Rep 2021;70(34):1156–62. doi: 10.15585/mmwr.mm7034e2 [published Online First: 2021/08/27]

35. Ortiz M. Texas nurses overwhelmed as ICU beds reach capacity https://spectrumlocalnews.com/tx/south-texas-el-paso/news/2021/08/05/texas-nurses-overwhelmed-as-icu-beds-reach-capacity-: Sectrum News, August 5 2021.

36. Teegardin C. As COVID-19 surge continues, Georgia hospitals running out of ICU beds. https://www.ajc.com/news/coronavirus/as-covid-19-surge-continues-georgia-hospitals-running-out-of-icu-beds/LQN7LSZNU5H7TADFLCYDKNZ2R4/: Atlanta Journal-Constitution, August 18 2021.

37. Muoio D. 10 states nearing—or exceeding—hospital capacity during COVID’s summer resurgence. https://www.fiercehealthcare.com/hospitals/10-states-nearing-or-exceeding-hospital-capacity-during-covid-s-summer-resurgence: Fierce Healthcare, August 19 2021.

38. Harris JE. COVID-19 Incidence and Hospitalization Rates are Inversely Related to Vaccination Coverage Among the 112 Most Populous Counties in the United States. https://www.medrxiv.org/content/10.1101/2021.08.17.21262195v2: medRxiv, August 20 2021.

